# Effectiveness of the Covid-19 vaccines in preventing infection in dental practitioners – results of a cross-sectional ‘questionnaire-based’ survey

**DOI:** 10.1101/2021.05.28.21257967

**Authors:** Sanjeev Kumar, Susmita Saxena, Mansi Atri, Sunil Kumar Chamola

## Abstract

India started its vaccination program at the beginning of 2021, the main beneficiaries being health workers and frontline workers including police, paramilitary forces, sanitation workers, and disaster management volunteers in the first phase. By the time, the second wave of Covid-19 impacted India, approximately 14 million healthcare and frontline workers, including dentists had been vaccinated.

**Aim:** To study the effectiveness of vaccination on a subset of high-risk healthcare workers i.e. dentists in preventing Covid-19 during the second wave of the pandemic.

**Study design:** A questionnaire based pan-India online survey was carried out to record the Covid-related experiences of dentists prior to and after vaccination.

**Result:** The sample size for this survey was 4493 respondents from across India. During the second wave, 9.18% (n=364) respondents became positive in spite of the vaccine, while 14.69%(n=78) became positive in the unvaccinated group. A chi-square test of independence was performed to examine the relation between vaccination and the Covid positivity rate in all age groups. The relation between these variables was highly significant, [*X2 (1, N = 4493) = 15*.*9809, p=*.*000064*].

**Conclusion:** Our pan-India online survey inferred that vaccination has a definitive role to play in reducing the positivity rate amongst dentists during the second wave of the pandemic across all age groups.

## Introduction

In early January 2021, the Drug Controller General of India (DCGI) approved two vaccines for clinical use, the Oxford–AstraZeneca vaccine, which is manufactured by the Serum Institute of India (SII) under the trade name “Covishield” and Bharat Biotech’s “Covaxin”.

On 16 January 2021, India began its massive vaccination program, targeting health workers and frontline workers including police, paramilitary forces, sanitation workers, and disaster management volunteers in the first phase. By 1 March 2021, 14 million healthcare and frontline workers, including dentists had been vaccinated.^1^ The next phase of the vaccine rollout covered all residents over the age of 60, residents between the ages of 45 and 60 with one or more qualifying comorbidities, and any health-care or frontline worker that did not receive a dose during phase 1.^2^

India is currently experiencing the onslaught of an aggressive second wave of Covid-19 infection which began around 11 February 2021 in Maharashtra. The situation progressively worsened in April 2021, ending the month by repeatedly setting new global records for daily cases. Two strains; the highly contagious U.K. variant (B.1.1.7) and the Indian variant(B.1.617) are believed to be causing this rapid rise in cases during the second wave.^3^ The overall nationwide test positivity rate was above 20% in April and the second wave was expected to peak around mid-May with infections exceeding 0.4 million/day.^4^ Till date, India has crossed 288,232 reported deaths^5^, of which more than 48,000 people have lost their lives to the disease in April 2021 alone.^3^

Dentistry is at the top of all professions most susceptible to contract Covid-19 because of the physical proximity to their patients.^6^ A spate of deaths due to Covid-19 within the dental community during the second wave has created anxiety among practitioners that the vaccines may not be as protective against the new mutant strains of SARS-CoV-2. No study has yet proven that the currently available vaccines in India are protective against the new variants in high-risk professions such as dentistry.

This cross-sectional questionnaire based pan-India online survey was carried out to record the Covid-related experiences of dentists prior to and after vaccination. The primary aim of the study was to assess whether vaccination has helped to reduce the positivity rate amongst dentists during the second wave.

The secondary objectives were to assess whether:

1. the vaccine protects against infection across all age groups?
2. the vaccine leads to less severe symptoms in vaccinated persons?
3. a single dose of vaccine is as effective as the two dose regime in preventing infection?
4. two dose regime of vaccine help to prevent more severe symptoms in persons compared to those who received only a single dose of vaccine?
5. vaccine is effective in preventing Covid 19 in persons with comorbidities?
6. Covishield is more effective than Covaxin in preventing infection or vice versa?

## Methodology

A cross-sectional survey was conducted from 29^th^ April to 2^nd^ May 2021, 11 weeks after the second wave of Covid-19 hit India. The data was collected online via a simple questionnaire of 15 items created on Google forms (annexure). Care was taken not to collect any information which would disclose the respondents’ identity and participation was completely voluntary.

The link for the Google Form was shared electronically in various dentistry related groups on Whatsapp, a freeware, cross-platform centralized messaging and voice-over-IP (VoIP) service owned by Facebook, Inc. The purpose for which the data was being collected was clearly specified in the accompanying message and the willingness to participate in the survey was considered as implied consent.

All responses received in the stipulated time-period were converted online to a Google spreadsheet for data analysis.

The data was analyzed online on https://www.socscistatistics.com/tests/. The output of the calculators and tools featured on this website has been audited for accuracy against the output produced by a number of established statistics packages, including SPSS and Minitab.^1^ The test applied was Chi square test of independence and the significance level was set at .05. The results were significant when *p*<.05.

## Results

From 29^th^ April to 2^nd^ May 2021, a total of 4493 responses were collected online. Out of 28 States and 8 Union Territories of India, responses were received from 26 states and 7 Union Territories (Table 1). No dentists responded from the states of Manipur and Nagaland and the UT of Ladakh. The age distribution of the respondents is shown in Graph 1. 74.43% (n=3344) respondents were below the age of 45 years, while 25.57%(n=1149) were above 45 years. 51.56% (n=2317) of the respondents were females while 47.96%(n=2155) were males. 21(0.46%) respondents did not reveal their sex.

**Graph 1:**
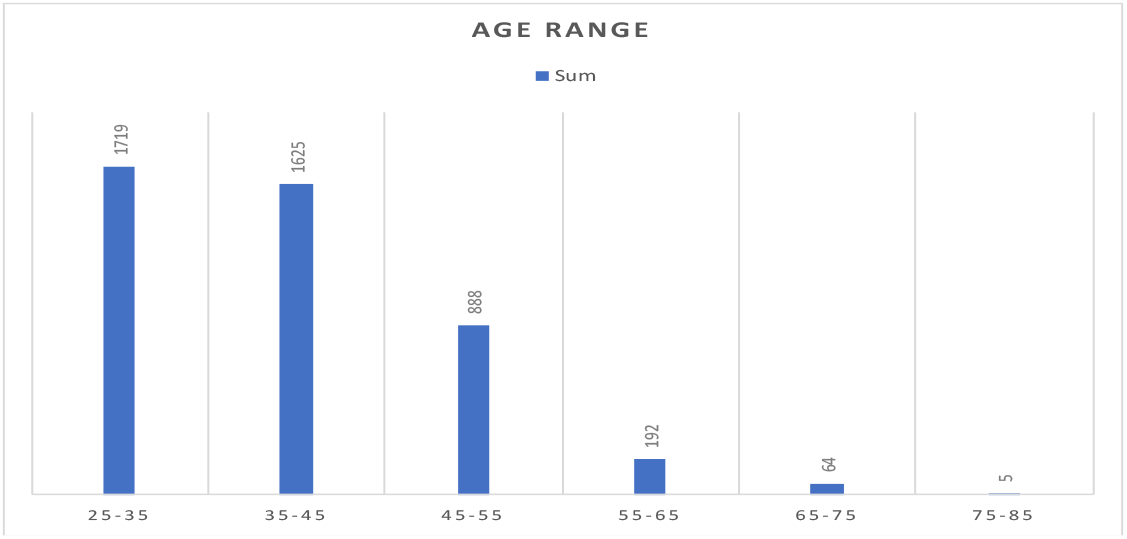
Age-wise distribution of respondents

**Table 1:** State-wise distribution of respondents

|  |  |  |
| --- | --- | --- |
| 1 | Maharashtra | 1495 |
| 2 | Karnataka | 587 |
| 3 | Gujarat | 367 |
| 4 | Uttar Pradesh | 322 |
| 5 | Andhra Pradesh | 265 |
| 6 | Telangana | 215 |
| 7 | Delhi | 200 |
| 8 | West Bengal | 149 |
| 9 | Punjab | 135 |
| 10 | Haryana | 97 |
| 11 | Tamil Nadu | 95 |
| 12 | Kerala | 93 |
| 13 | Tripura | 89 |
| 14 | Rajasthan | 82 |
| 15 | Madhya Pradesh | 49 |
| 16 | Jharkhand | 47 |
| 17 | Chandigarh | 41 |
| 18 | Chhattisgarh | 33 |
| 19 | Bihar | 22 |
| 20 | Himachal Pradesh | 19 |
| 21 | Uttarakhand | 16 |
| 22 | Sikkim | 14 |
| 23 | Jammu and Kashmir | 13 |
| 24 | Puducherry | 10 |
| 25 | Odisha | 10 |
| 26 | Goa | 9 |
| 27 | Assam | 8 |
| 28 | Meghalaya | 4 |
| 29 | Arunachal Pradesh | 2 |
| 30 | Lakshadweep | 2 |
| 31 | Mizoram | 1 |
| 32 | Andaman and Nicobar Islands | 1 |
| 33 | Dadra and Nagar Haveli | 1 |

81.08%(n=3643) were healthy individuals while 18.92% (n=850) had co-morbidities. Among those above 45 years of age, 36.98%(n=425) had associated co-morbidities, while 12.70%(n=425) among those below 45 years of age reported associated co-morbidities. This difference was statistically significant [*X2 (1, N = 4493) = 328*.*6389, p=*.*00001*].

In the age group of the respondents above 45 years having co-morbidities, 39.76%(n=169) were diabetic, while in the age group below 45 years having comorbidities, 24.24%(n=103) were diabetic. This difference was also statistically significant [*X2 (1, N =850) = 23*.*551, p=*.*00001*].(Table 2)

**Table 2:** showing the age-wise distribution of comorbidities and diabetes

|  | Co-morbidities | No Co-morbidities | Marginal Row Totals |
| --- | --- | --- | --- |
| **Below 45** | 425 (632.63) [68.14] | 2919 (2711.37) [15.9] | 3344 |
| **Over 45** | 425 (217.37) [198.32] | 724 (931.63) [46.27] | 1149 |
| **Marginal Column Totals** | 850 | 3643 | 4493 (Grand Total) |
| The chi-square statistic is 328.6389. The $p$-value is $< 0.00001$. Significant at $p < .05$. | | | |
|  | Diabetes | Other comorbidities | Marginal Row Totals |
| **Below 45** | 103 (136) [8.01] | 322 (289) [3.77] | 425 |
| **Above 45** | 169 (136) [8.01] | 256 (289) [3.77] | 425 |
| **Marginal Column Totals** | 272 | 578 | 850 (Grand Total) |
| The chi-square statistic is 23.551. The $p$-value is $< 0.00001$. Significant at $p < .05$. | | | |

During the vaccination drive, of 4493 respondents, 88.18%(n=3962) received at least one dose of either Covishield or Covaxin while 11.81%(n=531) did not receive vaccination. While 85.59%(n=2862) of the respondents below the age of 45 years were vaccinated, 95.73%(n=1100) above the age of 45 years were vaccinated. The difference in vaccination between the two age groups was statistically significant.(Table 3).

**Table 3:** Age-related difference in vaccinated vs non-vaccinated respondents

|  | Vaccinated | Not Vaccinated | Marginal Row Totals |
| --- | --- | --- | --- |
| **Below 45** | 2862 (2948.79) [2.55] | 482 (395.21) [19.06] | 3344 |
| **Over 45** | 1100 (1013.21) [7.43] | 49 (135.79) [55.47] | 1149 |
| **Marginal Column Totals** | 3962 | 531 | 4493 (Grand Total) |
| The chi-square statistic is 84.5251. The $p$-value is $< 0.00001$. Significant at $p < .05$. | | | |

During the second wave, 9.18% (n=364) respondents became positive in spite of the vaccine, while 14.69%(n=78) became positive in the unvaccinated group. A chi-square test of independence was performed to examine the relation between vaccination and the Covid positivity rate in all age groups. The relation between these variables was highly significant, [*X2 (1, N = 4493) = 15*.*9809, p=*.*000064*]. Age-related association between vaccination status and positivity rate was also studied. The relation between these variables for ages below 45 years was significant [*X2 (1, N = 3344) = 14*.*7204, p=*.*000125*], but for ages above 45 years [*X2 (1, N =1149) =0*.*1708, p=*.*679375*] it was not significant (Table 4).

**Table 4:** Age related difference in vaccination vs positivity rate

| All ages | Covid +ve | Covid-ve | Marginal Row Totals |
| --- | --- | --- | --- |
| **vaccinated** | 364 (389.76) [1.7] | 3598 (3572.24) [0.19] | 3962 |
| **nonvaccinated** | 78 (52.24) [12.71] | 453 (478.76) [1.39] | 531 |
| **Marginal Column Totals** | 442 | 4051 | 4493 (Grand Total) |
| The chi-square statistic is 15.9809. The $p$-value is .000064. Significant at $p < .05$. | | | |
|  | Covid +ve | Covid-ve | Marginal Row Totals |
| **<45 vaccinated** | 279 (302.97) [1.9] | 2583 (2559.03) [0.22] | 2862 |
| **<45 nonvaccinated** | 75 (51.03) [11.26] | 407 (430.97) [1.33] | 482 |
| **Marginal Column Totals** | 354 | 2990 | 3344 (Grand Total) |
| The chi-square statistic is 14.7204. The $p$-value is .000125. Significant at $p < .05$. | | | |
|  | Covid +ve | Covid-ve | Marginal Row Totals |
| **>45 vaccinated** | 85 (84.25) [0.01] | 1015 (1015.75) [0] | 1100 |
| **>45 nonvaccinated** | 3 (3.75) [0.15] | 46 (45.25) [0.01] | 49 |
| **Marginal Column Totals** | 88 | 1061 | 1149 (Grand Total) |
| The chi-square statistic is 0.1708. The $p$-value is .679375. Not significant at $p < .05$. | | | |
| Table 4: Age related difference in vaccination vs positivity rate |  |  |  |

At the time of the survey, 39.35%(n=1559) respondents were yet to complete their scheduled second vaccination while 60.25%(n=2387) had received both doses of the vaccine. 16 respondents did not mention the status of their vaccination. The data showed that 58.94 %(n=1681) below 45 years received both doses of the vaccine, as did 64.53%(n=706) above 45 years before the second wave hit India. This difference was significant statistically. (Table 5)

**Table 5:** Age related difference in incomplete(one) or completed (two doses) vaccination

|  | single dose | both doses | Marginal Row Totals |
| --- | --- | --- | --- |
| **Below 45** | 1171 (1126.78) [1.74] | 1681 (1725.22) [1.13] | 2852 |
| **Over 45** | 388 (432.22) [4.52] | 706 (661.78) [2.95] | 1094 |
| **Marginal Column Totals** | 1559 | 2387 | 3946 (Grand Total) |
| The chi-square statistic is 10.3484. The $p$-value is .001296. Significant at $p < .05$. | | | |

8.21%(n=128) respondents who had received just one dose, turned positive while 9.88%(n=236) respondents who received both doses, turned positive. This difference was however not statistically significant across all age groups [*X2 (1, N = 3946) =3*.*17, p=*.*075212*]. The positivity rate in respondents below 45 as well as in respondents above 45 years of age too did not differ significantly in those receiving single or both doses[*X2 (1, N = 2852) =3*.*7205, p=*.*053748*], [*X2 (1, N = 1094) =0*.*0732, p=*.*786696*]. (Table 6).

**Table 6:**
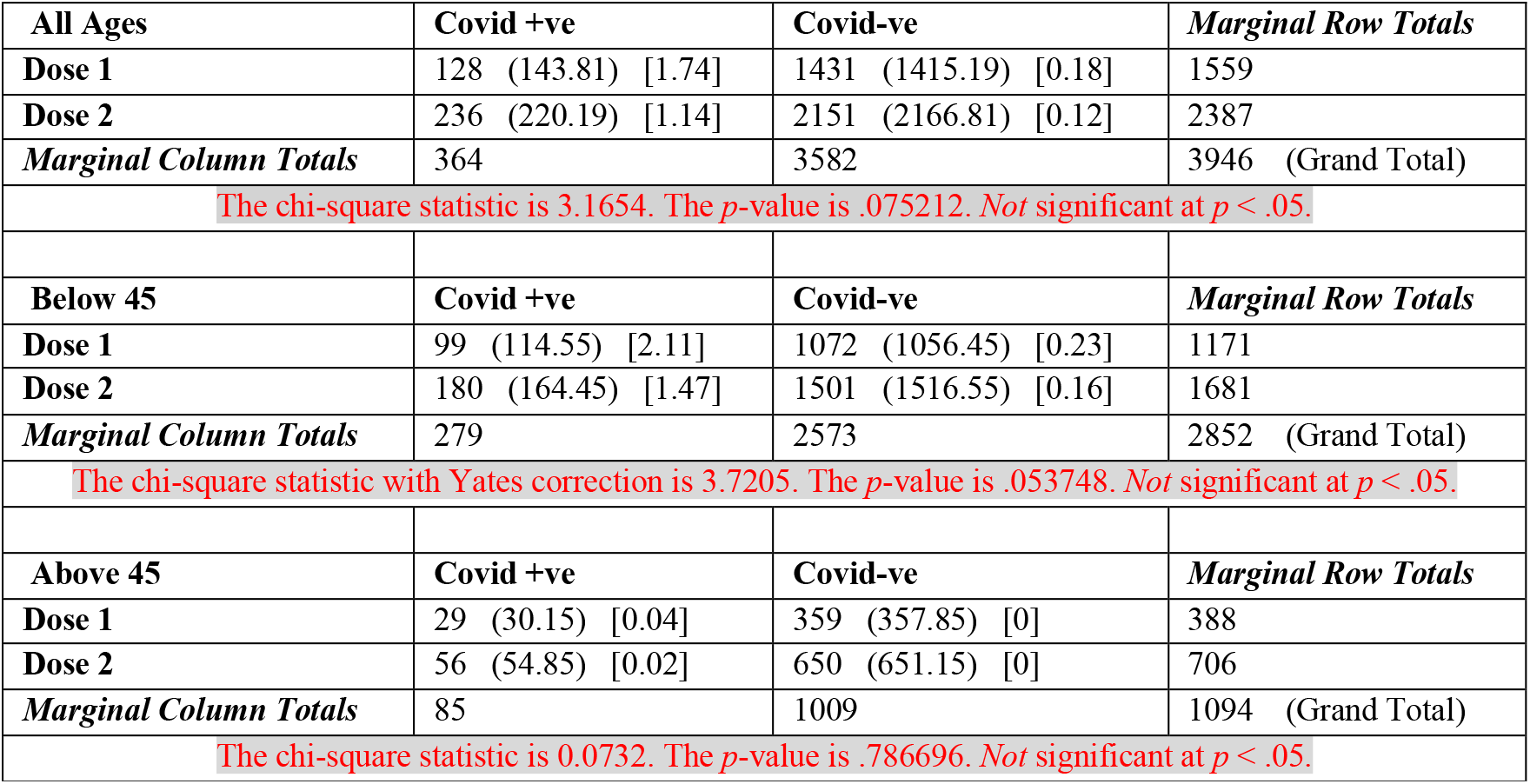
Single dose vs Two doses and positivity rate

In the unvaccinated group 71 of the 78 positive respondents described their symptoms. 78.87% (n=56) had mild and 21.13% (n=15) had moderate symptoms. In the vaccinated group, 348 of the 364 positive respondents their symptoms. 82.18%(n=288) described them as mild, 16.38%(n=57) as moderate and 1.44.%(n=5) as severe. Statistically, the difference between symptoms in vaccinated and non-vaccinated respondents was not significant [*X2 (1, N = 419) =0*.*4309, p=*.*511553*]. 79.68%(n=102) of the respondents who had only one dose of vaccination showed mild symptoms compared to 83.64% (n=184) who had both doses. Statistically, the difference between symptoms in those who received a single dose, or two doses was not significant [*X2 (1, N = 348) =0*.*8618, p=*.*35324*] (Table 7).

**Table 7:** Severity of symptoms in vaccinated vs non-vaccinated and single vs both doses recipients

|  | Mild | Moderate/Severe | Marginal Row Totals |
| --- | --- | --- | --- |
| vaccinated +ve | 286 (284.05) [0.01] | 62 (63.95) [0.06] | 348 |
| nonvaccinated +ve | 56 (57.95) [0.07] | 15 (13.05) [0.29] | 71 |
| Marginal Column Totals | 342 | 77 | 419 (Grand Total) |
| The chi-square statistic is 0.4309. The $p$-value is .511553. Not significant at $p < .05$. | | | |
|  | Mild | Moderate/Severe | Marginal Row Totals |
| Dose 1 | 102 (105.2) [0.1] | 26 (22.8) [0.45] | 128 |
| Dose 2 | 184 (180.8) [0.06] | 36 (39.2) [0.26] | 220 |
| Marginal Column Totals | 286 | 62 | 348 (Grand Total) |
| The chi-square statistic is 0.8618. The $p$-value is .35324. Not significant at $p < .05$. | | | |

In respondents with comorbidities, 8.4%(n=72) reported positive for Covid-19 compared to 10.16%(n=370) healthy respondents. This difference was not statistically significant [*X2 (1, N = 4493) =2*.*2084, p=*.*137259*]. 9.5% (n=303) of vaccinated respondents without comorbidities had Covid-19 compared to 7.9% (n=61) of vaccinated respondents with comorbidities. This difference was also not statistically significant [*X2 (1, N = 3962) =1*.*90, p=*.*168103*] 7.90%(n=61) vaccinated respondents with comorbidities tested positive compared to 14.10%(n=11) who did not receive the vaccine. This difference too was not statistically significant [*X2 (1, N = 850) =3*.*5135, p=*.*06087*] at p<.05(Table 8).

**Table 8:** Vaccination vs positivity rate in respondents with comorbidities

|  | Covid +ve | Covid-ve | Marginal Row Totals |
| --- | --- | --- | --- |
| without comorbidities | 370 (358.38) [0.38] | 3273 (3284.62) [0.04] | 3643 |
| with comorbidities | 72 (83.62) [1.61] | 778 (766.38) [0.18] | 850 |
| Marginal Column Totals | 442 | 4051 | 4493 (Grand Total) |
| The chi-square statistic is 2.2084. The $p$-value is .137259. Not significant at $p < .05$. | | | |
|  | Covid +ve | Covid-ve | Marginal Row Totals |
| without comorbidities+vaccinated | 303 (293.07) [0.34] | 2887 (2896.93) [0.03] | 3190 |
| with comorbidities+vaccinated | 61 (70.93) [1.39] | 711 (701.07) [0.14] | 772 |
| Marginal Column Totals | 364 | 3598 | 3962 (Grand Total) |
| The chi-square statistic is 1.8998. The $p$-value is .168103. Not significant at $p < .05$. | | | |
|  | Covid +ve | Covid-ve | Marginal Row Totals |
| with comorbidities+vaccinated | 61 (65.39) [0.3] | 711 (706.61) [0.03] | 772 |
| with comorbidities+nonvaccinated | 11 (6.61) [2.92] | 67 (71.39) [0.27] | 78 |
| Marginal Column Totals | 72 | 778 | 850 (Grand Total) |
| The chi-square statistic is 3.5135. The $p$-value is .06087. Not significant at $p < .05$. | | | |

Diabetic patients were analyzed separately because of their susceptibility to Covid-19. 32%(n=272) respondents with comorbidities were diabetic. Of these 94.85%(n=258) had received at least one dose of the vaccine. 7.36%(n=19) of these vaccinated respondents were infected with the virus. The chi square test for independence was not significant [*X2 (1, N = 272) =3*.*5333, p=*.*060146*] (Table 9).

**Table 9:**
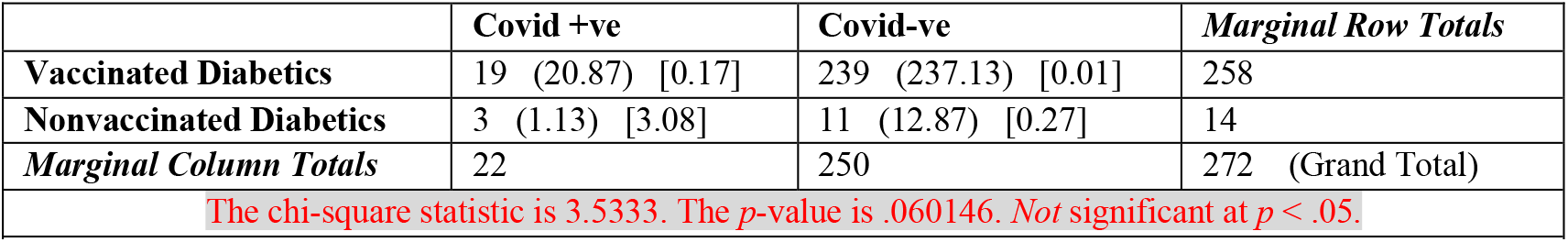
Vaccination vs positivity in respondents with diabetes

Among the total sample of 4493 respondents, 3962 (88.18%) had received at least one dose of vaccine. 3663(92.45%) had received the Covishield vaccine from Astra Zeneca, while 299(7.55%) had received Covaxin from Bharat Biotech. 8.36% respondents who received the Covaxin and 9.25% respondents who received Covishield turned positive during the second wave. Statistically, the difference between the two vaccines in their protective capability was not significant [*X2 (1, N = 3962) =0*.*26, p=*.*607033*] (Table 10).

**Table 10:**
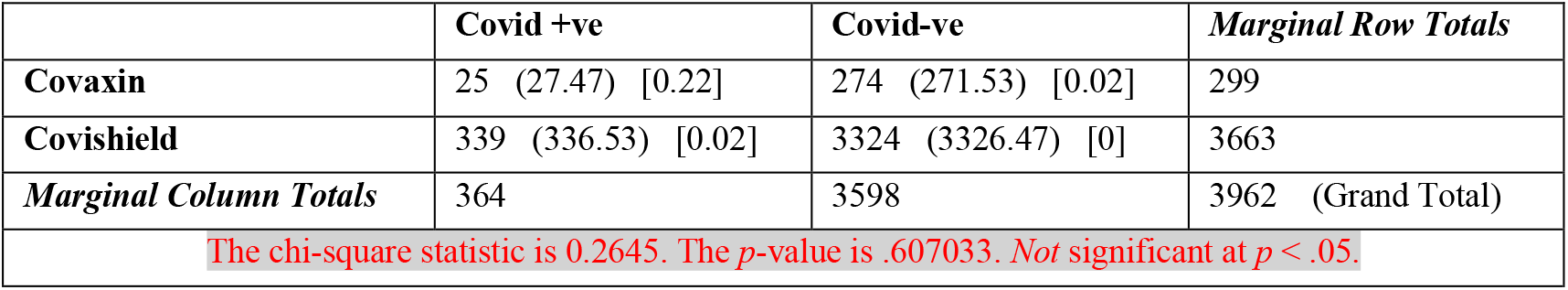
Effectivity of Covaxin vs Covishield in preventing infection

## Discussion

With a population of 1·38 billion, in the initial phase of the COVID-19 vaccination programme, India aimed at vaccinating 300 million people by August 2021, including 30 million health workers and frontline workers (eg, police, soldiers), and 270 million elderly people (ie, aged over 50 years) and people with co-morbidities. India’s strong domestic vaccine sector has enabled the country to launch one of the largest and fastest COVID-19 vaccination campaigns in the world.^7^

Two vaccines were launched simultaneously for clinical use in India: Covishield, developed by the Oxford-AstraZeneca and manufactured by the Serum Institute of India (SII) is prepared using the viral vector platform which utilizes a modified chimpanzee adenovirus – ChAdOx1 to carry the COVID-19 spike protein into the cells of humans and launch an immune response.^8^ The indigenous Covaxin has been developed by Hyderabad-based Bharat Biotech International Ltd in association with the Indian Council of Medical Research (ICMR) and the National Institute of Virology (NIV). It is developed by utilizing Whole-Virion Inactivated Vero Cell-derived technology.^8^

There is growing concern that emerging coronavirus variants may evade aspects of the immune responses stimulated by some vaccines. For example, in an analysis of about 2,000 people in South Africa, the AstraZeneca vaccine did not protect against mild or moderate COVID-19 due to the B.1.351variant.^9^ The current wave of Covid-19 in India was believed to be caused by several new variants especially U.K. variant (B.1.1.7) and the Indian variants (B.1.617 and B.1.618). By May 2021, B.1.617 variant has become the dominant strain across India and has spread to about 40 nations, including the United Kingdom, Fiji and Singapore prompting the World Health Organization to designate it as a ‘variant of concern’. Researchers from India have reported that some health-care workers in Delhi, vaccinated with Covishield had become reinfected with B.1.617. Reduction in the effectiveness of neutralizing antibodies generated by Covaxin against this variant, albeit small, have also been reported.^10^

Against this background, the present study was conducted to determine the effects of vaccination on positivity rate and severity of symptoms among Indian dentists during the second wave of Covid-19. Dentists were selected as the study sample as they are a high-risk sub-group of healthcare workers, in whom exposure to SARS-CoV-2 virus is likely to be more than in the general population. It is assumed that positive effects of the vaccine in this sub-group, if any, would be applicable to the whole population. The cue to analyze the data in two age groups separately i.e. above and below the age of 45 was taken from the Indian government’s vaccination strategy based on susceptibility to infection. In this study, almost 2/3^rd^ respondents were below the age of 45 indicative of the young dental workforce of India. Expectedly, the older age group had a significantly greater proportion of respondents with comorbidities including diabetes.

In our sample, 88.18% dentists participated in the government’s vaccination drive indicating a highly informed sub-group of healthcare workers. With over 95% compliance, the older age group of dentists showed exceptional enthusiasm in getting themselves vaccinated. In contrast, according to media reports less than 50% of frontline health workers had got vaccinated by 19^th^ April 2021.^11^

As a clear indication of efficacy of COVID-19 vaccines, our data indicates that non-vaccinated individuals across all age groups were more likely to get infected with the virus. The result corroborate the findings of similar studies done in other countries.^12,13^ Interestingly, according to our analysis, these vaccines seem to be more protective in the younger age group rather than in older individuals. The reason for this discrepancy may be a result of sampling error.

Several studies show that a high proportion of elderly individuals have suboptimal neutralising antibody responses following first dose vaccination with mRNA-based vaccines Pfizer-BioNTech and Moderna vaccine.^14,15,16,17^ Our data revealed that a single dose of the two vaccines available in India was as effective as two doses of vaccines in preventing infection in all age groups, including in the older age group of above 45 years. Since Covishield and Covaxin are both non-mRNA based vaccines, it is possible that the difference in formulation may be responsible for the increased immune response seen by even a single dose.

It has been reported that the COVID-19 vaccines help reduce severity of infection and need for hospitalization.^18^ This claim was refuted by our study, as no significant difference was observed in the severity of symptoms in vaccinated and non-vaccinated group or even in those receiving either a single or both doses of vaccine.

The novel coronavirus disease (COVID-19) tends to portend a poor prognosis in patients with diabetes mellitus (DM). Primary prevention with timely vaccination remains the mainstay for mitigating the risks associated with COVID-19 in patients with DM.^19^ In our sample, individuals with comorbidities did not show an increased susceptibility to infection compared to those without any comorbidities. Our analysis indicates that vaccines appear to be as effective in reducing the incidence of Covid-19 in the population with comorbidities as in healthy individuals. However, vaccines do not seem to reduce the positivity rate within this compromised group. Specifically, in respondents suffering from diabetes mellitus, the positivity rate did not change significantly with vaccination.

On-going research has shown that both Covaxin and Covishield are likely to be protective to limit the severity and mortality of the disease in the vaccinated individuals, in-spite of reduction in the neutralizing titer against B.1.617.1 variant.^20,21^ Our cross-sectional data demonstrated that currently there is little to choose between Covaxin and Covishield, both having similar capability to prevent Covid-19.

## Conclusion

Our pan-India online survey inferred that vaccination has a definitive role to play in reducing the positivity rate amongst dentists during the second wave across all age groups. In addition, a single of vaccine seems to be as effective as two doses of vaccine in preventing infection with SARS-CoV-2 virus. However, the vaccines did not reduce the severity of infection in the sample studied. The vaccines seem to be equally effective against infection in persons with co-morbidities as in healthy individuals. The two vaccines available currently in India, Covishield and Covaxin, are equally effective and there is no relation between the type of vaccine administered and the positivity rate among vaccinated persons.

## Data Availability

There is no external or supplementary material online at other repositories that pertain to this manuscript.

## Questionnaire

* Required

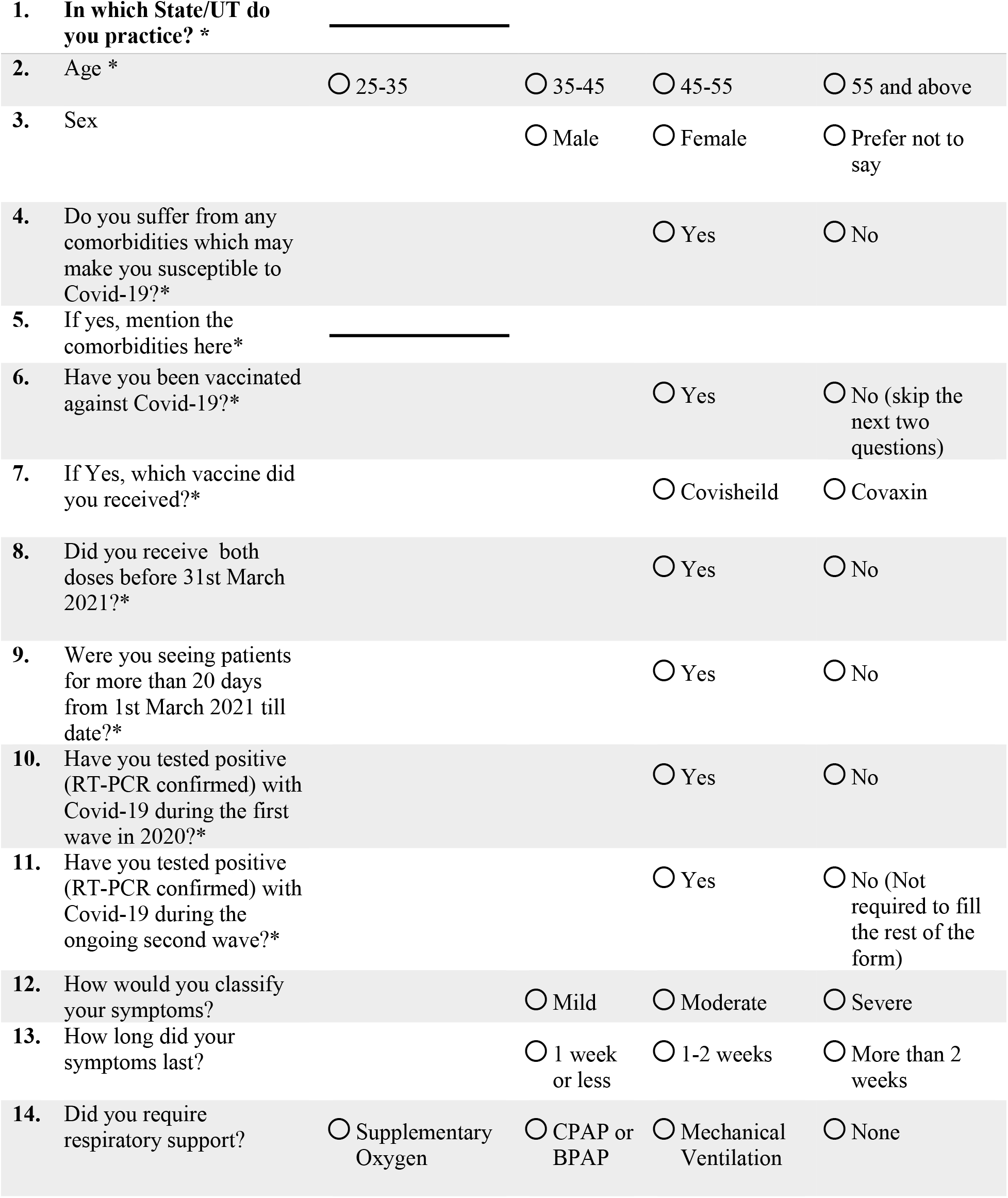
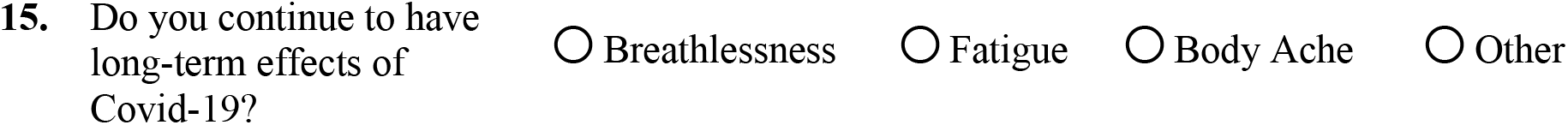

